# CMR Reveals Myocardial Damage from Cardiotoxic Oncologic Therapies in Breast Cancer Patients

**DOI:** 10.1101/2023.04.21.23288954

**Authors:** Johannes Kersten, Visnja Fink, Maria Kersten, Lisa May, Samuel Nunn, Marijana Tadic, Jens Huober, Inga Bekes, Michael Radermacher, Vinzenz Hombach, Wolfgang Rottbauer, Dominik Buckert

## Abstract

**Background:** Breast cancer is a common and increasingly treatable disease. However, survivors have a significantly elevated risk of cardiac events afterwards. This study aimed to characterise cardiac changes during cardiotoxic cancer therapy using cardiovascular magnetic resonance (CMR) imaging.

**Methods:** This study involved 34 patients with histologically proven breast cancer and planned cardiotoxic therapy. All patients underwent CMR before starting therapy, and 6 and 12 months thereafter. The CMR protocol included volumetric and functional analyses, parametric mapping, and deformation analysis using feature tracking. As the control group, 10 healthy female volunteers were scanned using the same protocol.

**Results:** With therapy, there was a significant reduction of left ventricular and right ventricular ejection fractions (both p < 0.05). Left ventricular radial (p = 0.008), circumferential (p = 0.010), and longitudinal strain (p = 0.036) were also reduced at follow-up. In the parametric mapping, there was a significant increase in native T1 time (1037 ± 41 ms vs. 1068 ± 51 ms vs. 1017 ± 57 ms, p < .001) and T2 time (55 ± 4 ms vs. 59 ± 3 ms vs. 57 ± 3 ms, p = 0.001), with unchanged extracellular volume and relative late gadolinium enhancement. Twelve months after cancer diagnosis, the breast cancer patients exhibited significant impairments in left ventricular global radial (p = 0.001), circumferential (p = 0.001), and longitudinal strain (p = 0.002) and T2 time (p = 0.008) compared to the healthy controls.

**Discussion:** Breast cancer patients receiving cardiotoxic chemotherapy show persistent deterioration in left ventricular strain values. This is accompanied by inflammatory changes in non-invasive tissue characterisation. Larger studies with longer follow-up periods are needed to identify patients at risk and establish preventive and therapeutic approaches.

## Introduction

Nearly one in two people develops cancer during their lifetime. The ageing population of developed countries has engendered a marked increase in this trend. Approximately 5% of the population survives after the diagnosis of a malignant disease (1), and patients are surviving cancer with increasing frequency (2). Therefore, research has turned to the side effects and long-term consequences of cancer therapies. In patients with Hodgkin’s lymphoma or breast cancer, the risk of cardiovascular complications exceeds that of the original tumour disease a few years after cancer therapy is administered (3,4). These complications can occur throughout life (5). Early detection or even prevention of these complications is the goal of current research.

Breast cancer is the most common malignancy in women, with approximately 2.1 million new cases worldwide each year (6). With a five-year survival rate of approximately 90%, it is a highly treatable affliction; this has been made possible largely by major research efforts (7). Therapeutic options for breast cancer include radiotherapy, chemotherapy, Her2-targeted therapy, anti-hormonal drugs, and various novel approaches, including the use of checkpoint inhibitors, in addition to surgical procedures. Many of these therapeutic options are potentially cardiotoxic.

Cardiovascular magnetic resonance (CMR) imaging, with its capacity for non-invasive tissue characterisation, as well as the structural and functional assessment of the heart, offers a method for better understanding cardiotoxic adverse events. Parametric mapping has become a key diagnostic tool for several cardiomyopathies and inflammatory changes (8–10). It could conceivably be used for the early detection of cardiotoxic side effects.

This study aimed to phenotype the structural and functional changes in the heart early after therapy application in patients with curable breast cancer.

## Methods

In this prospective single-centre longitudinal study, consecutive patients with newly diagnosed breast cancer were screened for enrolment. The inclusion criteria were histologic confirmation and planned cardiotoxic therapy in adjuvant or neoadjuvant intention, while the exclusion criteria were age under 18 years, evidence of metastases (M1-status), past surgery without complete removal of the tumour (R1 or R2), pregnancy, or pre-existing cardiovascular disease in the patient’s medical history. In addition, contraindications to CMR (e.g., claustrophobia, ferromagnetic implants) or CMR contrast media (e.g., known intolerances, glomerular filtration rate below 30 ml/min) led to exclusion.

The investigation plan included three CMR investigations: before the first therapy application, 6 months after therapy initiation, and 12 months after therapy initiation. Ten healthy women without clinical or anamnestic evidence of existing or past cardiovascular or oncological diseases were used as a control cohort and scanned once with the same CMR protocol.

The study was approved by the ethics committee of the University of Ulm (approval number 195/19) and conducted in accordance with the principles of the Declaration of Helsinki. Patients, controls or the public were not involved in the design, or conduct, or reporting, or dissemination plans of our research.

### Cardiovascular magnetic resonance

CMR was performed on all subjects using a 1.5T scanner with a 32-channel phased array cardiac surface coil (Achieva, Philips, Best, Netherlands). For volumetric and functional analyses, a balanced steady-state free-precision cine sequence (repetition time: 3.4 ms; echo time: 1.7 ms; slice thickness: 8 mm; no interslice gap; acquisition in end-expiration breath-hold) was used in three long-axis views (two-, three-, and four-chamber views) and contiguous short-axis views. The study protocol included a modified look-locker sequence in a 5(3)3 scheme for T1 mapping before and after the application of gadoterate meglumine (Dotarem®, Guerbet, Villepinte, France) in a dose of 0.2 mmol/kg of body weight. The T2 maps were obtained using a gradient spin-echo sequence. Late gadolinium enhancement (LGE) images (repetition time: 7.1 ms; echo time: 3.2 ms; slice thickness: 8 mm; respiratory navigator) were obtained 10 min after the administration of the contrast agent after individual adjustment for inversion time using a look-locker sequence.

All images were analysed by two experienced examiners in consensus using commercially available software (cvi42, Circle, Calgary, Canada). Left and right ventricular volumetry and myocardial mass were evaluated while excluding the papillary muscles using Simpson’s method, and the ejection fractions were calculated correspondingly. The deformation parameters were determined using two-dimensional feature-tracking analysis. Myocardial T1 and T2 values were obtained after contouring the endo- and epicardial borders in the corresponding sequences. Extracellular volume (ECV) maps were generated by the software after delineating the myocardium and a blood sample in native and post-contrast T1 maps using a standardised haematocrit of 0.4. ECV values were obtained analogously to T1 and T2 values. LGE was evaluated semi-quantitatively as the relative enhanced myocardium with a signal intensity above the five-fold standard deviation of a reference myocardium defined by the examiners.

### Statistics

For the descriptive analysis, continuous variables were expressed as means ± standard deviations, and categorical values were expressed as numbers and percentages. All data were normally distributed in a Kolmogorov–Smirnov test or graph. Ordinally scaled variables were compared using a two-sided Student’s t-test. In the case of more than two groups, an analysis of variance (ANOVA) was used, with the supplementary application of a post hoc test if necessary (Bonferroni). For nominally distributed variables, a chi-square test was used. To evaluate the possible factors influencing the development of cardiac dysfunction under tumour therapy, a univariate logistic regression analysis was performed. For every test, a two-tailed p-value of < 0.05 was considered statistically significant. The analysis was performed using IBM SPSS Statistics 26 (IBM, Armonk, NY, USA).

## Results

### Cohort and baseline characteristics

From December 2019 to August 2021, a total of 34 patients were enrolled in the study; one patient each was lost to follow-up after the first and the second CMR. The patients had a mean age of 50.2 ± 10.3 years, with a range of 33 to 71 years. Only a few cardiovascular risk factors were present: hypertension in 9/34 (26.5%) patients, diabetes mellitus in 3/34 (8.8%) patients, and dyslipidaemia and smoking each in 3/34 (8.8%) patients. Anthracycline-based chemotherapy regimens were predominantly used (91.2%), with a neoadjuvant approach in 79.4% and radiotherapy following chemotherapy in 76.5% of cases. The baseline characteristics, tumour biology, and cancer therapies used can be seen in Table 1. Four patients (11.8%) had previously been treated for cancer. None of the patients had a history of cardiovascular disease, and they had a normal cardiac biomarker at baseline, with a mean high-sensitivity troponin T of 3.7 ± 1.4 ng/l (norm < 15 ng/l) and N-terminal pro-brain natriuretic peptide of 73 ± 46 pg/ml (norm < 250 pg/ml).

**Table 1.**
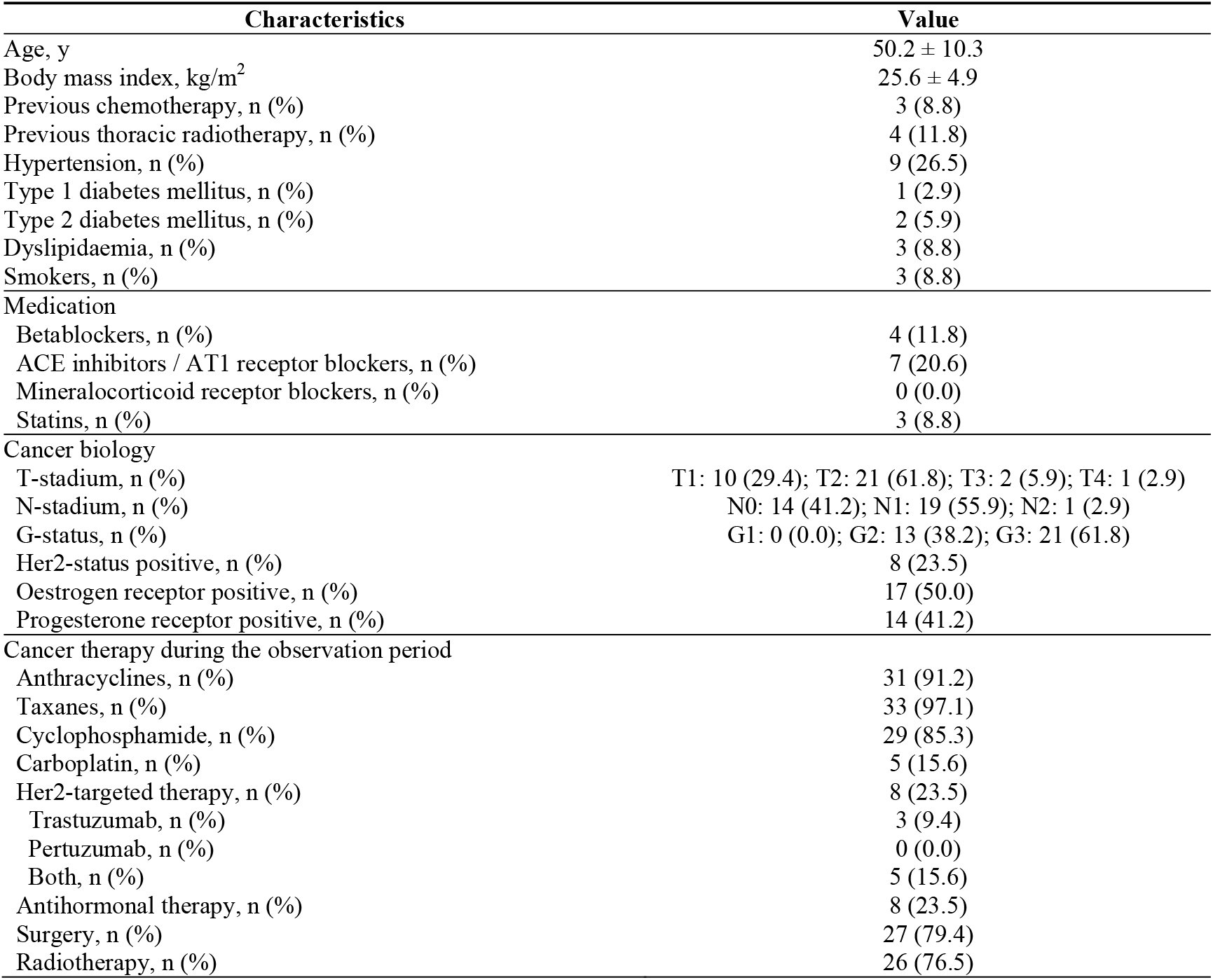
Patient characteristics (n = 34)

### Longitudinal results of repeated cardiovascular magnetic resonance

Prior to the initiation of cardiotoxic therapy, left and right ventricular volumes and functions were normal in all patients. Under therapy, there was a significant deterioration in the left ventricular ejection fraction (LVEF) from 65.2 ± 6.8% to 61.2 ± 6.7% at 6 months and 61.1 ± 6.1% at 12 months (p = 0.016). The decrease from baseline was also significant in the Bonferroni post hoc test at both follow-ups (p = 0.040 and p = 0.038, respectively). The lowest measured LVEF was 46.8%. The right ventricular ejection fraction (RVEF) also decreased significantly (59.9 ± 7.6% vs. 55.1 ± 6.2% vs. 56.7 ± 6.8%, p = 0.019). This was significant in the post hoc test when comparing the baseline measurement to the 6-month follow-up (p = 0.017), but not at 12 months (p = 0.204).

The deformation analysis showed a significant decrease in left ventricular strain values in all three orientations. The largest decrease was in global radial strain, with a relative decrease of 13.1% from 29.7 ± 6.1% to 26.5 ± 5.4% and 25.8 ± 4.1% (p = 0.008). This was significant both at 6 months (p = 0.044) and 12 months (p = 0.012) in the Bonferroni test. An example of a patient with worsening deformation parameters is illustrated in Figure 2.

Non-invasive tissue characterisation did not indicate acute or ongoing inflammatory cardiomyopathy prior to therapy. At 6 months, the parametric mapping showed a statistically significant but transient increase in native T1 (1037 ± 41 ms vs. 1068 ± 51 ms vs. 1017 ± 57 ms, p < .001) and T2 (55 ± 4 ms vs. 59 ± 3 ms vs. 57 ± 3 ms, p = 0.001). ECV and LGE showed no significant changes. An example of a native T1 and an ECV map 6 months after therapy initiation with a diffuse increase in T1 time is shown in Figure 3.

The volumetric and functional values and tissue characterisation values, as well as the respective time course, are shown in Table 2. The main results and changes in the CMR measurements are illustrated in Figure 1.

**Table 2.** Results of cardiovascular magnetic resonance imaging over time before cardiotoxic therapy (baseline), 6 months after therapy initiation, and 12 months after therapy initiation

| Value | Baseline<br>(n = 34) | After 6 months<br>(n = 33) | After 12 months<br>(n = 32) | p-value |
| --- | --- | --- | --- | --- |
| **Volumetry** |  |  |  |  |
| Left ventricular end-diastolic volume, ml | 127.2 ± 24.6 | 128.2 ± 23.1 | 127.6 ± 25.0 | 0.985 |
| Left ventricular end-diastolic volume indexed, ml/m<sup>2</sup> | 70.6 ± 12.1 | 71.0 ± 10.6 | 76.6 ± 14.2 | 0.096 |
| Left ventricular end-systolic volume, ml | 45.0 ± 14.6 | 50.3 ± 14.2 | 49.8 ± 12.9 | 0.230 |
| Left ventricular stroke volume, ml | 82.2 ± 15.1 | 77.9 ± 14.1 | 77.8 ± 17.0 | 0.413 |
| Left ventricular ejection fraction, % | 65.2 ± 6.8 | 61.2 ± 6.7 | 61.1 ± 6.1 | 0.016 |
| Left ventricular mass, g | 85.4 ± 17.9 | 95.9 ± 19.0 | 91.7 ± 17.6 | 0.064 |
| Right ventricular end-diastolic volume, ml | 129.6 ± 23.8 | 125.4 ± 25.3 | 132.5 ± 28.5 | 0.539 |
| Right ventricular end-diastolic volume indexed, ml/m<sup>2</sup> | 72.0 ± 11.7 | 69.7 ± 13.2 | 79.6 ± 16.7 | 0.013 |
| Right ventricular ejection fraction, % | 59.9 ± 7.6 | 55.1 ± 6.2 | 56.7 ± 6.8 | 0.019 |
| **Deformation parameters** |  |  |  |  |
| Left ventricular global radial strain, % | 29.7 ± 6.1 | 26.5 ± 5.4 | 25.8 ± 4.1 | 0.008 |
| Left ventricular global circumferential strain, % | -17.7 ± 2.2 | -16.5 ± 2.2 | -16.3 ± 1.8 | 0.010 |
| Left ventricular global longitudinal strain, % | -14.9 ± 2.1 | -13.6 ± 1.9 | -13.8 ± 2.5 | 0.036 |
| Right ventricular global longitudinal strain, % | -20.8 ± 4.5 | -21.1 ± 3.9 | -22.3 ± 3.7 | 0.318 |
| Left ventricular peak systolic radial strain rate, /sec | 1.6 ± 0.4 | 1.5 ± 0.3 | 1.4 ± 0.3 | 0.056 |
| Left ventricular peak systolic circumferential strain rate, /sec | -0.9 ± 0.4 | -1.0 ± 0.2 | -0.9 ± 0.2 | 0.481 |
| Left ventricular peak systolic longitudinal strain rate, /sec | -0.8 ± 0.2 | -0.8 ± 0.4 | -0.8 ± 0.2 | 0.880 |
| Right ventricular peak systolic longitudinal strain rate, /sec | -1.1 ± 1.0 | -1.1 ± 1.0 | -1.2 ± 0.6 | 0.844 |
| Left ventricular peak diastolic radial strain rate, /sec | -1.6 ± 0.5 | -1.3 ± 0.8 | -1.3 ± 0.3 | 0.041 |
| Left ventricular peak diastolic circumferential strain rate, /sec | 0.9 ± 0.2 | 0.9 ± 0.2 | 0.8 ± 0.1 | 0.024 |
| Left ventricular peak diastolic longitudinal strain rate, /sec | 0.8 ± 0.3 | 0.8 ± 0.2 | 0.7 ± 0.2 | 0.058 |
| Right ventricular peak diastolic longitudinal strain rate, /sec | 1.4 ± 0.5 | 1.3 ± 0.6 | 1.3 ± 0.5 | 0.627 |
| **Tissue characterisation** |  |  |  |  |
| T1 native, ms | 1037 ± 41 | 1068 ± 51 | 1017 ± 57 | < .001 |
| T2, ms | 55 ± 4 | 59 ± 3 | 57 ± 3 | 0.001 |
| Extracellular volume | 25.1 ± 3.0 | 25.9 ± 3.3 | 25.9 ± 3.2 | 0.464 |
| Late gadolinium enhancement (relative), % | 2.4 ± 3.9 | 3.5 ± 3.4 | 4.0 ± 5.8 | 0.332 |

**Figure 1.**
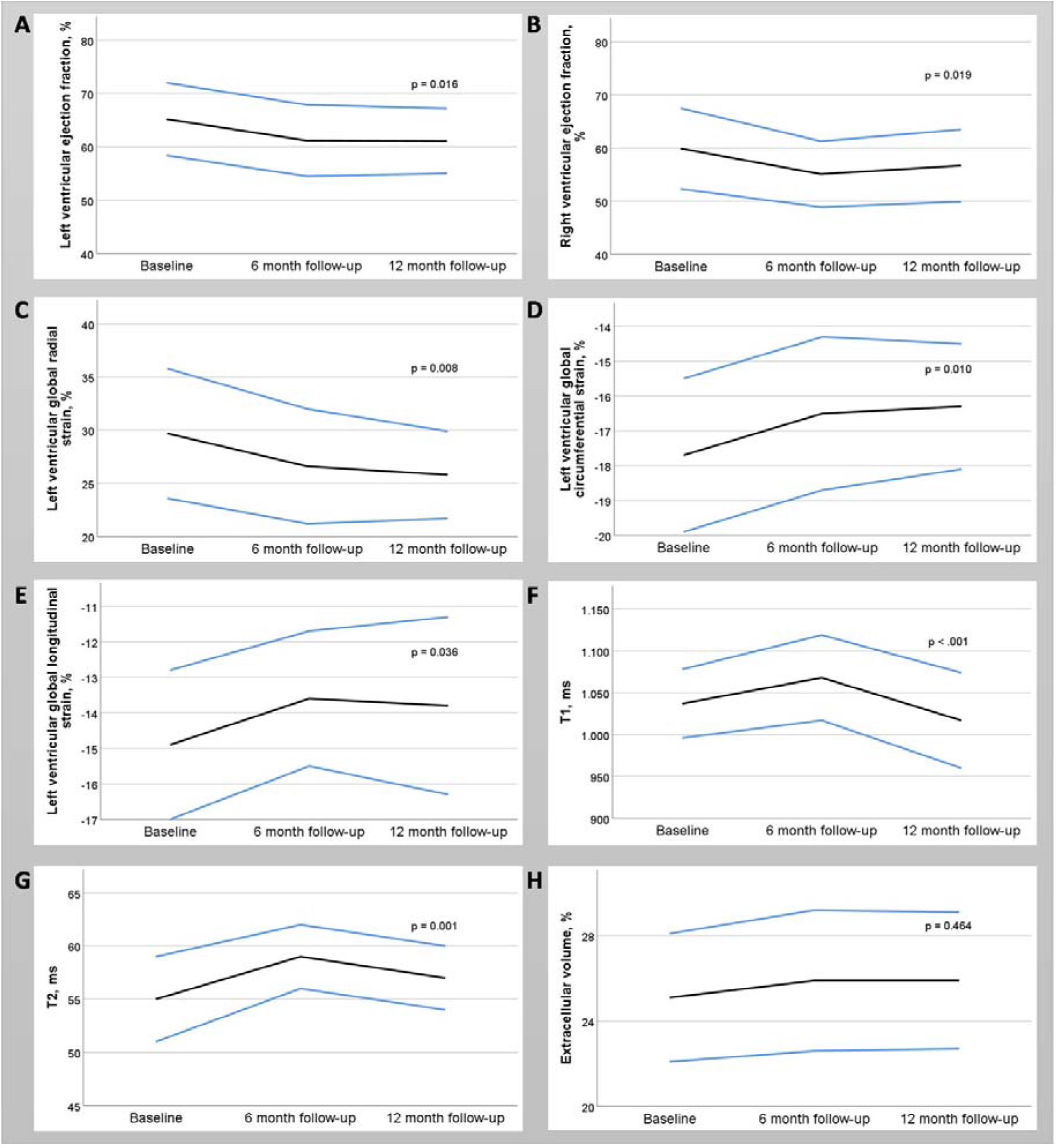
The major results of cardiovascular magnetic resonance imaging of patients undergoing cardiotoxic therapy for breast cancer. The changes in the mean values (black lines) over time and one-fold standard deviations (blue lines) are shown. There is a significant decrease in left (A) and right (B) ventricular ejection fractions. The deformation analysis shows significant deterioration of radial (C), circumferential (D), and longitudinal strain (E) in the left ventricle. Right ventricular longitudinal strain showed no significant changes (not illustrated). The parametric mapping shows a transient increase in native T1 and T2 times, while extracellular volume remains unchanged.

**Figure 2.**
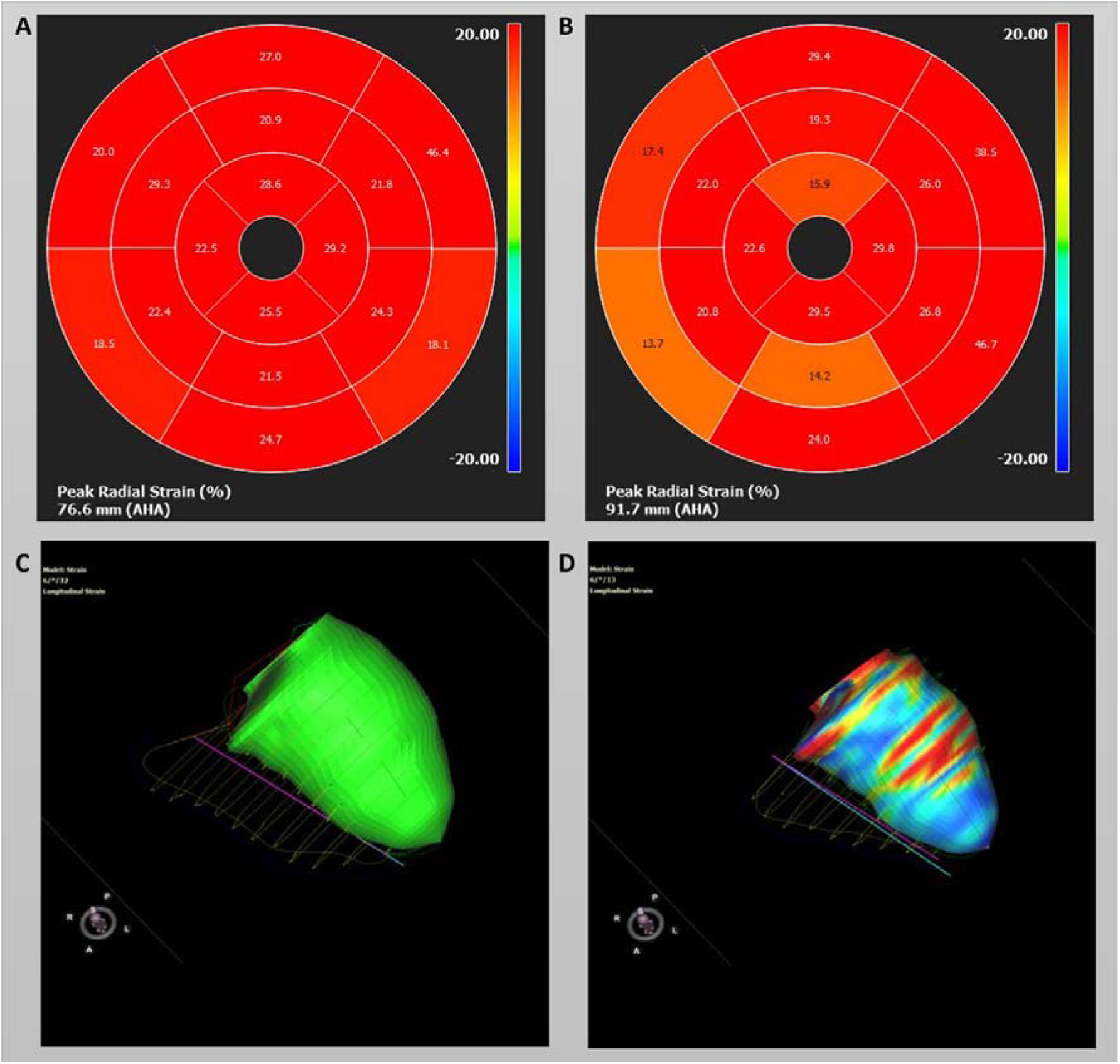
Strain analysis using feature tracking in a patient undergoing cardiotoxic therapy. Left ventricular global radial strain showed diffuse deterioration at the 6-month follow-up (B) in an initially (A) completely normal 16-segment model. In addition, a deterioration of left ventricular global longitudinal strain was also observed after 6 months. A colour-coded 3D visualisation in end-diastolic (C) and end-systolic (D) phases is shown. The end-systolic phase would have been expected to have a homogeneous blue colouration under normal contraction.

**Figure 3.**
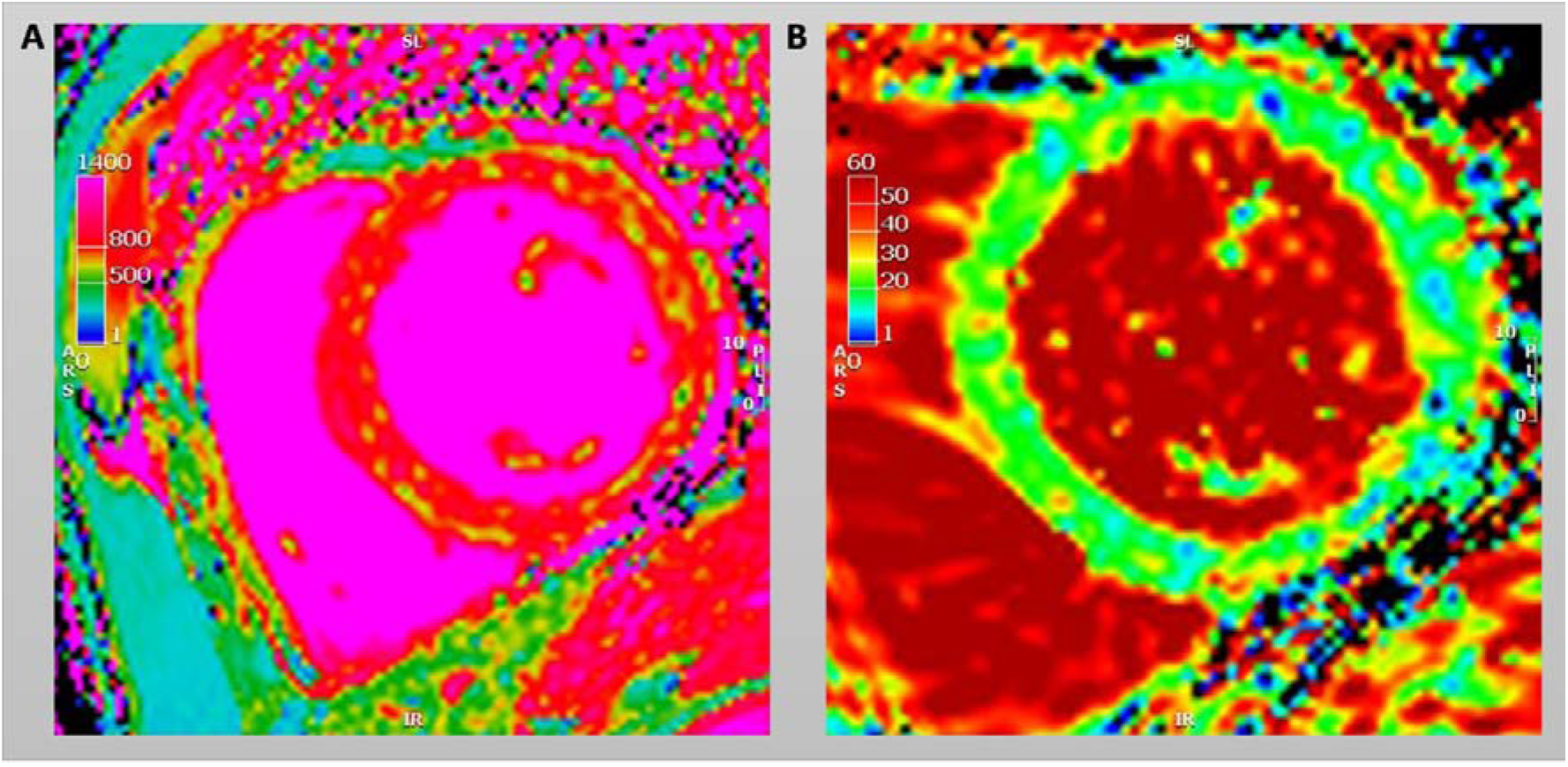
An example of a native T1 (A) and an ECV map (B) of the patient in Figure 2. The examination was performed at the 6-month follow-up.

Furthermore, 18/32 (56.3%) patients met the ESC guideline definition of at least mild cancer treatment-related cardiac dysfunction (CTRCD), while 3/31 (9.4%) patients met that of moderate CTRCD. The patients with and without CTRCD were not significantly different in terms of age, body mass index, Her2 status, or long-term medication. The patients with CTRCD were all treated with a combination of anthracycline and taxane, supplemented in four cases by Her2-targeted therapy (trastuzumab alone or in combination with pertuzumab). A total of 13 patients (72.2%) received radiotherapy during the study period; this was also not different between the patients with and without CTRCD. Moreover, no differences were found between the two groups in the other therapies. In the logistic regression analysis, no associations were found for patient characteristics or types of therapy with the occurrence of CTRCD (see also Table S1 in the supplementary material).

### Comparison with healthy controls

Ten female subjects were recruited as controls. They were significantly younger than the patients (41.3 ± 14.6 years vs. 50.2 ± 10.3 years, p = 0.034); were not taking any long-term medication; and had no prior history of cardiovascular or malignant diseases. Compared to the patients, the healthy controls had equal values for left and right ventricular volumetry and global function (all p > 0.05). In particular, LVEF and RVEF were not different (LVEF: 61.1 ± 6.1% vs. 63.4 ± 5.4%, p = 0.296 and RVEF: 56.8 ± 6.8% vs. 57.5 ± 6.6%, p = 0.746). There were already significant differences between the longitudinal strain values of the patients and the controls at baseline (p = 0.005), but not for the radial (p = 0.094) or circumferential strain (p = 0.083). The absolute difference was considerably more pronounced after cardiotoxic therapy. All left ventricular strain values were reduced in the patient cohort at the 12-month follow-up, including radial (25.8 ± 4.1% vs. 33.0 ± 8.3%, p = 0.001), circumferential (−16.3 ± 1.8% vs. -18.9 ± 2.8%, p = 0.001), and longitudinal strain (−13.8 ± 2.5% vs. -16.6 ± 2.1%, p = 0.002). Furthermore, left ventricular diastolic strain rates were significantly reduced in the patients with breast cancer, as Table 3 shows. The parametric mapping revealed an increase in T2 values in the patient cohort (57 ± 3 ms vs. 54 ± 4 ms, p = 0.008). The remaining parameters of tissue characterisation were equal to those of the control group.

**Table 3.** Cardiac magnetic resonance imaging of all patients 12 months after therapy initiation in comparison to healthy female controls (n = 10).

| Value | Patients after 12<br>months | Healthy females | p-value |
| --- | --- | --- | --- |
| **Volumetry** |  |  |  |
| Left ventricular end-diastolic volume, ml | 127.6 ± 25.0 | 138.5 ± 32.7 | 0.271 |
| Left ventricular end-diastolic volume indexed, ml/m<sup>2</sup> | 76.6 ± 14.2 | 76.5 ± 17.5 | 0.974 |
| Left ventricular end-systolic volume, ml | 49.8 ± 12.9 | 51.5 ± 17.1 | 0.735 |
| Left ventricular stroke volume, ml | 77.8 ± 17.0 | 87.0 ± 18.7 | 0.155 |
| Left ventricular ejection fraction, % | 61.1 ± 6.1 | 63.4 ± 5.4 | 0.296 |
| Left ventricular mass, g | 91.7 ± 17.6 | 93.2 ± 15.9 | 0.814 |
| Right ventricular end-diastolic volume, ml | 132.5 ± 28.5 | 143.6 ± 28.6 | 0.291 |
| Right ventricular end-diastolic volume indexed, ml/m<sup>2</sup> | 79.6 ± 16.7 | 79.2 ± 14.3 | 0.947 |
| Right ventricular ejection fraction, % | 56.8 ± 6.8 | 57.5 ± 6.6 | 0.746 |
| **Deformation parameters** |  |  |  |
| Left ventricular global radial strain, % | 25.8 ± 4.1 | 33.0 ± 8.3 | 0.001 |
| Left ventricular global circumferential strain, % | -16.3 ± 1.8 | -18.9 ± 2.8 | 0.001 |
| Left ventricular global longitudinal strain, % | -13.8 ± 2.5 | -16.6 ± 2.1 | 0.002 |
| Right ventricular global longitudinal strain, % | -22.3 ± 3.7 | -21.9 ± 4.1 | 0.817 |
| Left ventricular peak systolic radial strain rate, /sec | 1.4 ± 0.3 | 1.7 ± 0.5 | 0.006 |
| Left ventricular peak systolic circumferential strain rate, /sec | -0.9 ± 0.2 | -1.0 ± 0.2 | 0.332 |
| Left ventricular peak systolic longitudinal strain rate, /sec | -0.8 ± 0.2 | -0.9 ± 0.2 | 0.173 |
| Right ventricular peak systolic longitudinal strain rate, /sec | -1.2 ± 0.6 | -1.6 ± 0.5 | 0.142 |
| Left ventricular peak diastolic radial strain rate, /sec | -1.3 ± 0.3 | -1.6 ± 0.3 | 0.005 |
| Left ventricular peak diastolic circumferential strain rate, /sec | 0.8 ± 0.1 | 0.9 ± 0.2 | 0.042 |
| Left ventricular peak diastolic longitudinal strain rate, /sec | 0.7 ± 0.2 | 0.9 ± 0.3 | 0.039 |
| Right ventricular peak diastolic longitudinal strain rate, /sec | 1.3 ± 0.5 | 1.3 ± 0.3 | 0.821 |
| **Tissue characterisation** |  |  |  |
| T1 native, ms | $1017 \pm 57$ | $1028 \pm 23$ | 0.572 |
| T2, ms | $57 \pm 3$ | $54 \pm 4$ | 0.008 |
| Extracellular volume | $25.9 \pm 3.2$ | $23.7 \pm 2.8$ | 0.063 |
| Late gadolinium enhancement (relative), % | $4.0 \pm 5.8$ | $2.0 \pm 2.0$ | 0.306 |

## Discussion

The main findings of the present study are as follows: (1) Cardiotoxic therapy in breast cancer results in changes in non-invasive tissue characterisation that are comparable to those of inflammatory cardiomyopathies, with a transient increase in native T1 and T2. (2) Significant reductions in left ventricular function and deformation parameters occur with cancer therapy, with no predictive factors allowing risk stratification. (3) Changes in cardiac function are still detectable in breast cancer patients after 12 months using deformation analysis compared to healthy controls.

Non-invasive tissue characterisation using parametric mapping and LGE imaging is a key component of myocardial inflammation diagnostics. According to the updated Lake–Louise criteria, a T2 elevation indicating oedema and a change in native T1, ECV, or LGE indicating myocardial damage are the primary criteria for the diagnosis of myocarditis (8). The secondary criteria include evidence of pericarditis and myocardial dysfunction. In our cohort, an increase in T2 and native T1 time that was at least transient, with relative myocardial dysfunction, was seen in a considerable proportion of the patients, indicating inflammation. Previous studies have found evidence of chemotherapy-induced myocarditis. In a study involving pigs, a regional increase in T2 time was demonstrated after intracoronary anthracycline injection, which recovered after cessation (11). In another study by Haslbauer et al., the patients underwent CMR either early (< 3 months) or late (> 12 months) after chemotherapy (12). In comparison to the healthy controls, the early cohort showed native T1 and T2 elevation (T1: 1137 ± 61 ms vs. 1053 ± 21 ms; T2: 50 ± 5 ms vs. 44 ± 3 ms), while the late cohort showed T1 elevation (1121 ± 47 ms vs. 1053 ± 21 ms) and left ventricular dysfunction (LVEF: 48 ± 12% vs. 60 ± 7; left ventricular global longitudinal strain: -17 ± 11% vs. -24 ± 5%). Based on these results, the authors hypothesised that myocardial inflammation leads to later myocardial dysfunction. A limiting factor was that no pre-therapeutic CMR was available for comparison, and the groups could not be considered a priori as longitudinal. The present study is relatively small, with 32 complete follow-ups, but it has good longitudinal assessability.

Other studies have also failed to find significant changes in LGE, similar to the present study. This does not seem surprising, as diffuse rather than localised changes are to be expected under cancer therapy (13,14).

In our cohort, there was a significant reduction in left ventricular longitudinal strain at the baseline itself. It has been previously shown that cancer patients can have subclinical changes in the deformation analysis even if they are therapy-naïve (15). Tadic et al.’s retrospective study using echocardiography examined the strain values in 122 cancer patients before the start of therapy and found a reduction in all three dimensions (16). The probability of systolic impairment is significantly higher in haematooncological diseases (17); however, humoral factors favouring myocardial dysfunction have also been discovered in breast cancer (18). Cancer causes the release of inflammatory cytokines. The extent of inflammation is prognostically influential for both cancer and cardiac health (17,19–21). Furthermore, as shown in the CANTOS trial, proinflammatory cytokines promote both cancerogenesis and cardiovascular events in the beginning (22,23). An existing pre-therapeutic change in non-invasive tissue characterisation in patients with cancer, such as an increase in T1 and T2, would also be conceivable. However, in our cohort, there were no differences between the healthy volunteers and the patients with regard to parametric mapping.

The equal LVEF in the cancer patients at 12 months and the healthy subjects, despite the otherwise significantly changed strain values, highlights the importance of deformation analysis. Thus, significantly more patients can be identified with CTRCD than with pure volumetric analysis, which is the reason behind ESC guidelines recommending left ventricular longitudinal strain as a monitoring tool (24). In other studies, the longitudinal strain has been shown to have prognostic importance (17,25). The majority of studies on deformation analysis in cancer employ speckle-tracking echocardiography. However, there is good comparability to CMR feature tracking, although there is no complete intermodal interchangeability (26,27). Compared to echocardiography, CMR offers the possibility of a one-stop-shop examination with excellent interobserver reproducibility and the possibility of non-invasive tissue characterisation. Future studies should examine whether this has further prognostic implications.

In the present study, the regression analysis showed no association between the baseline characteristics, such as cardiovascular risk factors, and the occurrence of CTRCD. Other studies have found an increase in the risk of cardiac dysfunction in older patients as well as in patients with obesity, hypertension, and diabetes mellitus. Furthermore, an increase was also observed depending on the therapy regime, such as with the combined administration of anthracyclines and Her2-targeted therapy or additive radiation (28,29). The present study is limited in this respect due to the limited number of cases. Investigating the influence of anthracycline-free chemotherapy on cardiovascular health in the long term, especially in breast cancer patients, may be of further interest (30).

## Conclusions

Potential cardiotoxic therapy in breast cancer patients is associated with changes in non-invasive tissue characterisation that are consistent with inflammatory processes. This is accompanied by functional changes, especially in deformation analysis, which remain persistent 12 months after therapy initiation. Over half of the patients had to be diagnosed with CTRCD; this raises further questions regarding surveillance and risk stratification, especially when considering the absolute frequency of the disease. This study also gives rise to the question of whether targeted drug or non-drug interventions can positively influence the structural and functional changes in the heart, namely with regard to imaging-guided cardioprotection.

## Limitations

The main limitation of this study is the monocentric design with a limited cohort size. Furthermore, long-term imaging and clinical data would be desirable, as cardiovascular events in cancer survivors often occur years or decades after therapy, especially with thoracic radiotherapy. Moreover, no biomarkers, such as troponin, were recorded in a standardised manner in our study at follow-up. However, these will certainly be part of the standard diagnostics in the surveillance of cardiotoxic therapy.

## Data Availability

The data that support the findings of this study are available from the corresponding author upon reasonable request.

## List of abbreviations

CMR: Cardiovascular magnetic resonance
CTRCD: Cancer therapy-related cardiac dysfunction
ECV: Extracellular volume
LGE: Late gadolinium enhancement
LVEF: Left ventricular ejection fraction
RVEF: Right ventricular ejection fraction

## References

1. Arndt V, Dahm S, Kraywinkel K. Krebsprävalenz in Deutschland 2017. Onkologe 2021;27(8):717–23.

2. Arnold M, Rutherford MJ, Bardot A, et al. Progress in cancer survival, mortality, and incidence in seven high-income countries 1995-2014 (ICBP SURVMARK-2): a population-based study. The Lancet. Oncology 2019;20(11):1493–505.

3. Lu Z, Teng Y, Ning X, Wang H, Feng W, Ou C. Long-term risk of cardiovascular disease mortality among classic Hodgkin lymphoma survivors. Cancer 2022.

4. Patnaik JL, Byers T, DiGuiseppi C, Dabelea D, Denberg TD. Cardiovascular disease competes with breast cancer as the leading cause of death for older females diagnosed with breast cancer: a retrospective cohort study. Breast cancer research BCR 2011;13(3):R64.

5. van Nimwegen FA, Schaapveld M, Janus CPM, et al. Cardiovascular disease after Hodgkin lymphoma treatment: 40-year disease risk. JAMA internal medicine 2015;175(6):1007–17.

6. Bray F, Ferlay J, Soerjomataram I, Siegel RL, Torre LA, Jemal A. Global cancer statistics 2018: GLOBOCAN estimates of incidence and mortality worldwide for 36 cancers in 185 countries. CA: a cancer journal for clinicians 2018;68(6):394–424.

7. Siegel RL, Miller KD, Fuchs HE, Jemal A. Cancer statistics, 2022. CA: a cancer journal for clinicians 2022;72(1):7–33.

8. Ferreira VM, Schulz-Menger J, Holmvang G, et al. Cardiovascular Magnetic Resonance in Nonischemic Myocardial Inflammation: Expert Recommendations. Journal of the American College of Cardiology 2018;72(24):3158–76.

9. Kersten J, Heck T, Tuchek L, Rottbauer W, Buckert D. The Role of Native T1 Mapping in the Diagnosis of Myocarditis in a Real-World Setting. Journal of clinical medicine 2020;9(12).

10. Haaf P, Garg P, Messroghli DR, Broadbent DA, Greenwood JP, Plein S. Cardiac T1 Mapping and Extracellular Volume (ECV) in clinical practice: a comprehensive review. Journal of cardiovascular magnetic resonance official journal of the Society for Cardiovascular Magnetic Resonance 2016;18(1):89.

11. Galán-Arriola C, Lobo M, Vílchez-Tschischke JP, et al. Serial Magnetic Resonance Imaging to Identify Early Stages of Anthracycline-Induced Cardiotoxicity. Journal of the American College of Cardiology 2019;73(7):779–91.

12. Haslbauer JD, Lindner S, Valbuena-Lopez S, et al. CMR imaging biosignature of cardiac involvement due to cancer-related treatment by T1 and T2 mapping. International journal of cardiology 2019;275:179–86.

13. Drafts BC, Twomley KM, D’Agostino R, et al. Low to moderate dose anthracycline-based chemotherapy is associated with early noninvasive imaging evidence of subclinical cardiovascular disease. JACC. Cardiovascular imaging 2013;6(8):877–85.

14. Toro-Salazar OH, Gillan E, O’Loughlin MT, et al. Occult cardiotoxicity in childhood cancer survivors exposed to anthracycline therapy. Circulation. Cardiovascular imaging 2013;6(6):873–80.

15. Fabiani I, Panichella G, Aimo A, et al. Subclinical cardiac damage in cancer patients before chemotherapy. Heart failure reviews 2022;27(4):1091–104.

16. Tadic M, Genger M, Baudisch A, et al. Left Ventricular Strain in Chemotherapy-Naive and Radiotherapy-Naive Patients With Cancer. The Canadian journal of cardiology 2018;34(3):281–7.

17. Kang Y, Assuncao BL, Denduluri S, et al. Symptomatic Heart Failure in Acute Leukemia Patients Treated With Anthracyclines. JACC. CardioOncology 2019;1(2):208–17.

18. Maayah ZH, Takahara S, Alam AS, et al. Breast cancer diagnosis is associated with relative left ventricular hypertrophy and elevated endothelin-1 signaling. BMC cancer 2020;20(1):751.

19. Kalogeropoulos AP, Georgiopoulou VV, Butler J. From risk factors to structural heart disease: the role of inflammation. Heart failure clinics 2012;8(1):113–23.

20. Proctor MJ, Morrison DS, Talwar D, et al. An inflammation-based prognostic score (mGPS) predicts cancer survival independent of tumour site: a Glasgow Inflammation Outcome Study. British journal of cancer 2011;104(4):726–34.

21. Mousavi N, Tan TC, Ali M, Halpern EF, Wang L, Scherrer-Crosbie M. Echocardiographic parameters of left ventricular size and function as predictors of symptomatic heart failure in patients with a left ventricular ejection fraction of 50-59% treated with anthracyclines. European heart journal. Cardiovascular Imaging 2015;16(9):977–84.

22. Ridker PM, Everett BM, Thuren T, et al. Antiinflammatory Therapy with Canakinumab for Atherosclerotic Disease. The New England journal of medicine 2017;377(12):1119–31.

23. Aday AW, Ridker PM. Antiinflammatory Therapy in Clinical Care: The CANTOS Trial and Beyond. Frontiers in cardiovascular medicine 2018;5:62.

24. Lyon AR, López-Fernández T, Couch LS, et al. 2022 ESC Guidelines on cardio-oncology developed in collaboration with the European Hematology Association (EHA), the European Society for Therapeutic Radiology and Oncology (ESTRO) and the International Cardio-Oncology Society (IC-OS). European heart journal 2022;43(41):4229–361.

25. Ali MT, Yucel E, Bouras S, et al. Myocardial Strain Is Associated with Adverse Clinical Cardiac Events in Patients Treated with Anthracyclines. Journal of the American Society of Echocardiography official publication of the American Society of Echocardiography 2016;29(6):522-527.e3.

26. Valente F, Gutierrez L, Rodríguez-Eyras L, et al. Cardiac magnetic resonance longitudinal strain analysis in acute ST-segment elevation myocardial infarction: A comparison with speckle-tracking echocardiography. International journal of cardiology. Heart & vasculature 2020;29:100560.

27. Pryds K, Larsen AH, Hansen MS, et al. Myocardial strain assessed by feature tracking cardiac magnetic resonance in patients with a variety of cardiovascular diseases - A comparison with echocardiography. Scientific reports 2019;9(1):11296.

28. McGowan JV, Chung R, Maulik A, Piotrowska I, Walker JM, Yellon DM. Anthracycline Chemotherapy and Cardiotoxicity. Cardiovascular Drugs and Therapy 2017;31(1):63–75.

29. Carver JR, Shapiro CL, Ng A, et al. American Society of Clinical Oncology clinical evidence review on the ongoing care of adult cancer survivors: cardiac and pulmonary late effects. Journal of clinical oncology official journal of the American Society of Clinical Oncology 2007;25(25):3991–4008.

30. Yu K-D, Liu X-Y, Chen L, et al. Anthracycline-free or short-term regimen as adjuvant chemotherapy for operable breast cancer: A phase III randomized non-inferiority trial. The Lancet regional health. Western Pacific 2021;11:100158.

